# Estimate of Covid prevalence using imperfect data

**DOI:** 10.1101/2020.04.14.20064840

**Authors:** Witold R. Rudnicki, Radosław Piliszek

## Abstract

The real number of people who were truly infected with SARS-CoV-2, is certainly significantly larger than the official record. Few countries have tracking and testing procedures that are sufficiently robust to discover nearly all infections. In most countries they are inadequate, hence the true extent of the pandemic is unknown. The current study proposes the estimate of the COVID-19 extent for countries with sufficiently high number of deaths and cases. The estimate is based on a simple model of mortality. This model was developed for a reference country with a large number of cases and high intensity of COVID-19 testing. The model is then applied to compute apparent mortality in the target and reference countries. The number of cases in the target country is then estimated assuming constant underlying true mortality. The estimate of cases in most countries is significantly higher than the official record. As of April 12, 2020, the global estimate is 5.2 million compared to 1.8 million in the official record. The models developed in this study are available at covid-model.net. The model ignores several factors that are known to influence mortality, such as the demographics and health condition of population, state of epidemic and sociological differences between countries. While the model is rough, it nevertheless provides a unified approach to producing a systematic global estimate of the extent of the COVID-19 epidemic and can be useful for its monitoring.

## 1 Introduction

A new pneumonia disease of viral origin, COVID-19, caused by SARS-CoV-2, appeared in China in December 2019 [1, 2]. The infection quickly spread worldwide [3]. As of April 12, 2020, the officially recorded number of cases reached 1,852,257 and the number of deaths officially attributed to COVID-19 reached 114,194^1^.

The true prevalence of the disease is certainly larger than the officially recorded cases. The differences in testing protocols, available resources and policies between countries result in widely varying estimates of mortality and prevalence of COVID [4]. Omitting the asymptomatic and mild cases from testing, results in a large apparent mortality and underestimation of the true number of infections.

On the other hand, the testing policy influences the number of deaths attributed to COVID-19 to a much lower extent. Some cases may be lost due to insufficient testing. Additionally, the policies of attributing deaths to COVID-19 or other co-occuring conditions may vary between countries. Nevertheless, the influence of these factors on the final death count should be lower than the influence of the testing procedures on the count of cases.

Therefore, one can use the official death count and the estimate of mortality to imply the number of cases. Unfortunately, the estimates of the mortality are highly variable and depend on the applied methodology [5, 6, 7, 8] as well as true fraction of asymptotic cases. The fraction of asymptomatic infections in COVID-19 has been estimated to be less than 20% using Diamond Princess data [9]. It has been also implicitly estimated using epidemics modelling to be more than 90% [10]. It is also known, that mortality varies strongly with age, comorbidity, and the ability of a healthcare system to cope with the epidemic [11, 12]. An additional problem for applying the mortality estimates from the countries in different phase of the epidemic, is that the distribution of times between onset of symptoms and death is known very imprecisely.

In the current study we propose a methodology that generates an estimate on the current state of epidemic in a given country using the reported deaths and reference epidemiology data from the reference country. The model aims at estimating what would be the number of positive tests, if testing was performed to the standards of the best testing countries. The reference country is selected using known testing policies, a sufficiently long cases history to develop models, and the state of the epidemic that is similar to target countries.

## 2 Models

The data for modelling COVID-19 epidemics is obtained from the John Hopkins University [13]. All models are based on the simple assumption, that the underlying biology of the viral infection is identical in all countries, and therefore the differences in the apparent mortality rate arise due to the different levels of detection of true cases.

This assumption is certainly an oversimplification, since it is already known that the mortality varies very strongly between age groups and therefore the true mortality in a given country depends on the population structure. What is more, the mortality depends on the capacity of the healthcare system in the country to provide the appropriate treatment to patients. This capacity may depend also on the state of the epidemic, and resources available to build the adequate temporary infrastructure etc. Another factor in variability in mortality can be attributed to differences in social behaviour - e.g. how prevalent are multi-generational households, the frequencies and intensity of social contacts, in particular between younger and older generations etc. The number of factors contributing to the true mortality is so large that it is impossible to account for them within the time-frame of the study. Therefore, all these factors are omitted and left to the country-based adjustment which can be done externally. Another assumption underlying the study is that the countries which are most likely to detect the highest percentage of asymptomatic and mild cases are those which perform most tests per capita and at the same time have low reported mortality. Hence, the estimate of true mortality using these countries is the least biased.

The total number of cases in the target country is obtained in all models as:

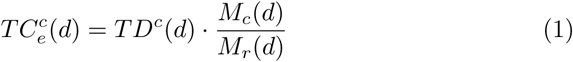

where, 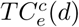 is an estimate of total number of cases in the target country, *M*_*c*_(*d*) is an apparent mortality in the target country and *M*_*r*_(*d*) is an apparent mortality in the reference country.

### 2.1 Model 1

The first model directly converts epidemic state in a reference country to the target country. The estimate of mortality of COVID-19 in a country for day *d* is obtained as

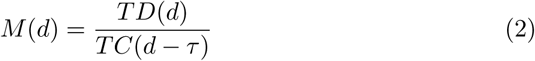

where, *M* (*d*) is an estimate of mortality at day *d, TD*(*d*) is a total number of deaths recorded at the day *d*, and *TC*(*d − τ*) is a total number of cases recorded *τ* days before day *d*. The *τ* is an empirically obtained parameter that corresponds to the delay between the time of test and the death of the patient. The value of the *τ* was determined empirically for the reference country for model 2.

### 2.2 Models 2 and 3

Both second and third model are simplifications of the following formal model of deaths:

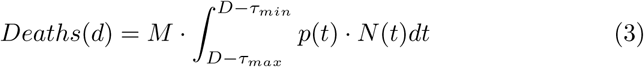

where number of new deaths in day *d* is obtained as the integral over time of the distribution of death times from the onset of symptoms, multiplied by a number of new cases in a given time. Stating this differently, when the true number of new cases on day *t* is given by *N* (*t*), then the number of deaths on day *t* + *τ* is determined by the overall mortality *M* and probability that death will occur in day *t* + *τ*.

**Model 2** uses the point estimate of the probability distribution *p*(*t*) from equation 3 in the following form:

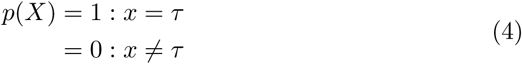

The mortality is then estimated from parameters of the simple linear regression without intercept, where current number of deaths is proportional to the current number of deaths:

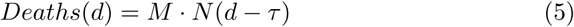

**Model 3** uses the uniform estimate of the probability distribution *p*(*t*) from equation 3 in the following form:

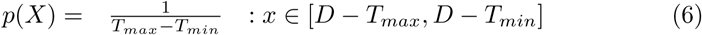

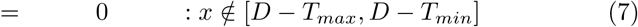

The mortality is then estimated from the average number of new cases in the period starting *d−T*_*max*_ day earlier and ending *d−T*_*min*_ days earlier, again using linear regression without the intercept:

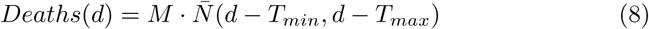

Both Model 2 and Model 3 are drastic simplifications of the model described by the Eq. 3. Nevertheless, in the exponential growth phase they have one significant advantage over more realistic distributions, namely they require fewer parameters, and therefore are less prone to overfitting. Effectively, they represent the effects of convolution of distribution of time to death and exponential growth. All parameters for the models, namely the time offset between test result and death *τ* and time span for deaths [*T*_*min*_, *T*_*max*_ were derived for the reference country by fitting models 2 and 3. The quality of the fit between the models and data is estimated using the square of the correlation coefficient between number of deaths estimated by the model and the true data. The final form of the models is then obtained as a weighted average with Model 1:

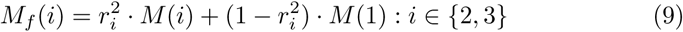

where *M*_*f*_ (*i*) is the final estimate of mortality for Model *i*, 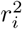 is the square of the correlation coefficient between deaths and earlier cases for Model *i, M* (1) is the estimate of mortality from Model 1, and *M* (*i*) for *i* ∈ {2, 3} is the regression coefficient from Equations 5 and 8, respectively.

### 2.3 Error estimates

The upper and lower estimate for Model 1, was obtained respectively as:

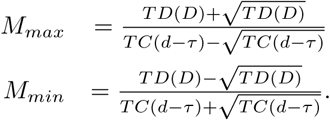

For Models 2 and 3, the standard deviation of parameters of linear regression were computed and the estimates of the upper and lower bounds for the model are obtained as:

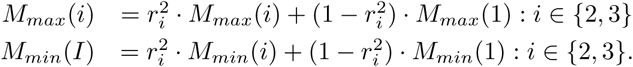

The final estimate of mortality in the target country is obtained by computing results of available models and taking the average of them. The number of available models depends on the available data. Models are computed only for countries that have reached at least 30 total deaths, a sufficiently long time before building of the model. The time span required is at least 3, 17 or 23 days, for models 1, 2 and 3, respectively. The estimates of the lower and upper bounds of the estimate are obtained by averaging the bounds computed for available models.

## 3 Reference Country

The reference was selected with the following requirements:

- the reference country should have a high level of testing,
- a healthcare system that hasn’t been overwhelmed with the epidemic,
- low mortality,
- a relatively long history of epidemics development,
- and a sufficiently large number of cases and deaths that would allow to build a reliable model.

An additional requirement for consideration is the level of political freedom that allows independent monitoring of the official statistics. There are two countries that fit this description best, namely South Korea and Germany, see Figure 1. Among this pair Germany has far more registered deaths (3,022 deaths registered in Germany and 214 in South Korea, as of April 12, 2020)^2^What is more, Germany is currently in the growth phase of the epidemic with nearly 4000 new cases recorded daily, whereas South Korea has managed to supress the epidemics, from over 500 daily cases in late February and early March to the level of less than 30 cases in April 10^2^.

**Figure 1:**
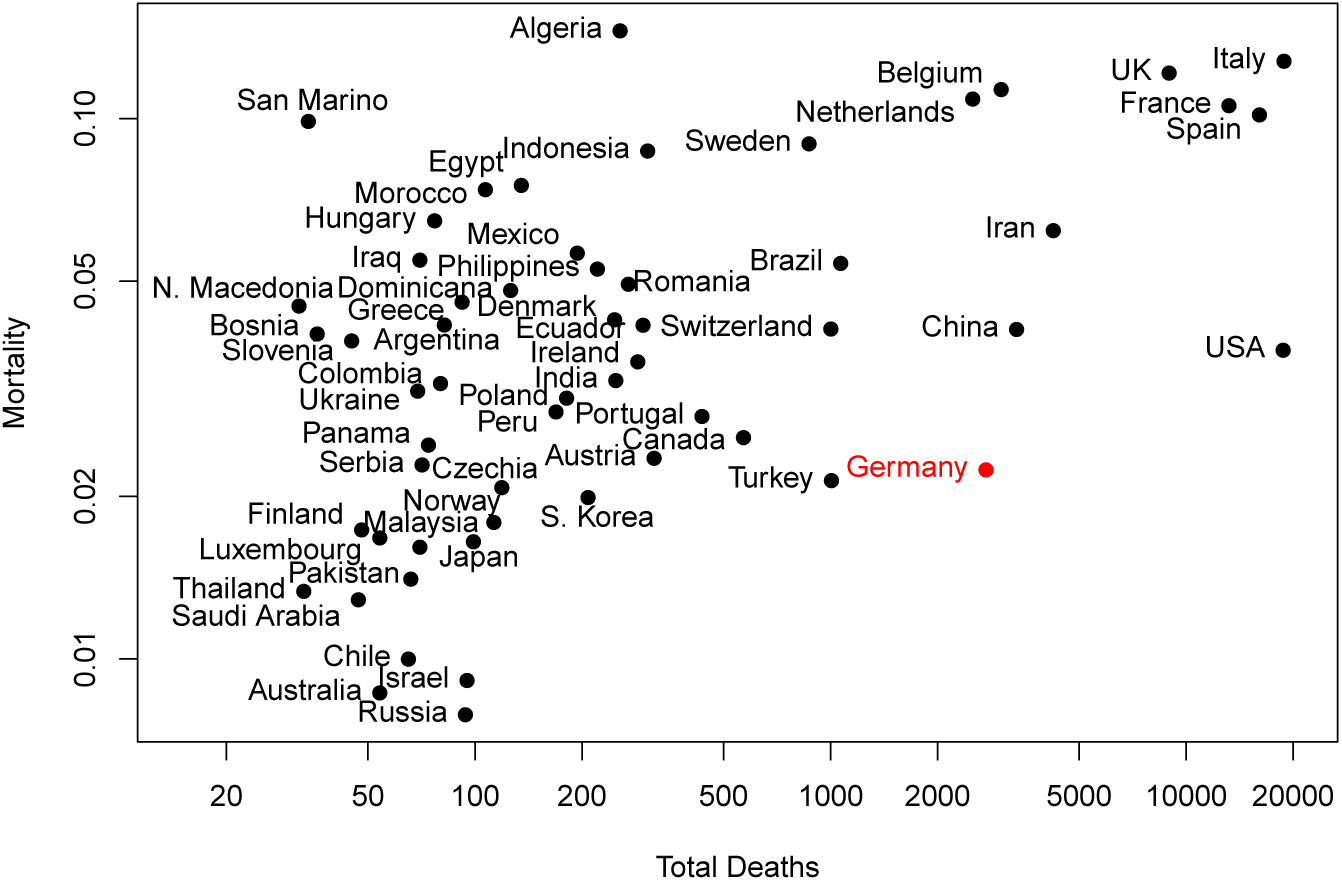
Simple estimation of Covid mortality for countries with more than thirty registered deaths. The mortality is computed by dividing the total number of deaths by total number of cases.

To examine the quality of the models for reference country, the modelling procedure was performed nine times with data terminated at each of the first nine days of April 2020. The values of mortalities, as well as the estimates of quality of fit of the model obtained for Germany, are shown in Table 1.

**Table 1:** Mortality and quality of fit for three models obtained for Germany. *M* ^1^, 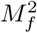 and 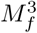 are final estimates of mortality obtained in Models 1, 2 and 3, respectively; *M* ^2^ and *M* ^3^ are estimates of mortality obtained from the linear regression fit in Models 2 and 3; *r*^2^(*M*_2_) and *r*^2^(*M*_3_) are r-squared computed for the linear regression fit in Models 2 and 3.

| Date | $M^1$ | $M^2$ | $r^2(M_2)$ | $M_f^2$ | $M^3$ | $r^2(M_3)$ | $M_f^3$ |
| --- | --- | --- | --- | --- | --- | --- | --- |
| 2020-04-01 | 0.0246 | 0.0257 | 0.74 | 0.0255 | 0.0321 | 0.99 | 0.0321 |
| 2020-04-02 | 0.0252 | 0.0264 | 0.82 | 0.0262 | 0.0338 | 0.98 | 0.0337 |
| 2020-04-03 | 0.0251 | 0.0259 | 0.85 | 0.0258 | 0.0341 | 0.98 | 0.0341 |
| 2020-04-04 | 0.0250 | 0.0257 | 0.87 | 0.0256 | 0.0341 | 0.98 | 0.0341 |
| 2020-04-05 | 0.0255 | 0.0261 | 0.86 | 0.0261 | 0.0330 | 0.93 | 0.0329 |
| 2020-04-06 | 0.0271 | 0.0278 | 0.79 | 0.0277 | 0.0342 | 0.92 | 0.0342 |
| 2020-04-07 | 0.0281 | 0.0288 | 0.77 | 0.0287 | 0.0344 | 0.93 | 0.0344 |
| 2020-04-08 | 0.0302 | 0.0315 | 0.71 | 0.0312 | 0.0372 | 0.85 | 0.0370 |
| 2020-04-09 | 0.0307 | 0.0322 | 0.74 | 0.0319 | 0.0364 | 0.87 | 0.0378 |

Models 1 and 2, give a similar estimate of mortality that is substantially lower than that of Model 3. This happens because the number of cases used by these models is recorded with a 7 days offset with the number of deaths, whereas in Model 3, the number of cases is average over time span that is significantly shifted towards longer times, effectively contributing less cases to the denominator in Eq. 2.

On the other hand, the Model 3 fits reference data to a much larger extent, with r-squared varying between 0.85 and nearly perfect 0.99. All models give a relatively stable estimate of mortality for the first five days, but then the estimates starts to rise systematically. What is more, the quality of the fit between the model and the observations also decreases after the first five days. There are several possible explanations for this effect. This drift may arise due to the changing dynamics of the epidemic, resulting in a worse fit of the models to the data.

The observed increase of apparent mortality may also signal that, at the end of March, the extent of testing became systematically worse relative to the number of true cases. The increase of mortality could also be genuine due to approaching the limits of healthcare system capacity, or due to a larger number of older patients infected later in the epidemic. It is likely that all factors contribute to some extent.

## 4 Results and Discussion

The models described in the previous sections were applied to all countries in the world for which data is available. The mortality obtained for countries for which all three models were computed is presented in Table 2. The correction factors derived from the models are displayed along the mortality. They are defined as:

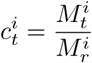

where, *C*^i^ is a correction factor obtained from model *i* for the target country *t*; 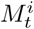 and 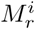 are the mortalities computed from model *i* for target and reference country, respectively.

**Table 2:** Mortality and quality of fit for three models for countries with largest number of recorded deaths. *M*^1^, 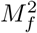 and 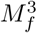 are estimates of mortality obtained in models 1, 2 and 3, respectively; *c*^1^, *c*^2^ and *c*^3^ are correction factors derived from models 1, 2 and 3; *r*^2^(*M*_2_) and *r*^2^(*M*_3_) are r-squared computed for the models 2 and 3. Countries are sorted in a increasing order by value of mortality computed from Model 1. Only countries for which all three models were computed are shown. All computations performed for data available at April 10.

| Country | $M^1$ | $c^1$ | $r^2(M_2)$ | $M_f^2$ | $c^2$ | $r^2(M_3)$ | $M_f^3$ | $c^3$ |
| --- | --- | --- | --- | --- | --- | --- | --- | --- |
| South Korea | 0.020 | 1.00 | 0.09 | 0.018 | 1.00 | 0.08 | 0.051 | 1.34 |
| Austria | 0.026 | 1.00 | 0.02 | 0.027 | 1.00 | 0.20 | 0.029 | 1.00 |
| Germany | 0.031 | 1.00 | 0.74 | 0.032 | 1.00 | 0.87 | 0.036 | 1.00 |
| China | 0.041 | 1.33 | 0.23 | 0.034 | 1.07 | 0.01 | 0.093 | 2.46 |
| Portugal | 0.045 | 1.47 | 0.70 | 0.041 | 1.28 | 0.26 | 0.048 | 1.28 |
| Turkey | 0.050 | 1.63 | 0.82 | 0.042 | 1.31 | 0.77 | 0.053 | 1.40 |
| Switzerland | 0.050 | 1.64 | 0.04 | 0.048 | 1.51 | 0.09 | 0.059 | 1.56 |
| USA | 0.069 | 2.25 | 0.96 | 0.068 | 2.12 | 0.96 | 0.081 | 2.16 |
| Iran | 0.081 | 2.65 | 0.74 | 0.081 | 2.26 | 0.01 | 0.074 | 1.95 |
| Brazil | 0.117 | 3.81 | 0.83 | 0.113 | 3.55 | 0.90 | 0.154 | 4.06 |
| France | 0.136 | 4.43 | 0.82 | 0.124 | 3.88 | 0.01 | 0.150 | 3.98 |
| Spain | 0.138 | 4.50 | 0.75 | 0.122 | 3.83 | 0.11 | 0.149 | 3.94 |
| Sweden | 0.142 | 4.63 | 0.67 | 0.174 | 5.46 | 0.71 | 0.221 | 5.84 |
| Italy | 0.159 | 5.17 | 0.86 | 0.148 | 4.65 | 0.01 | 0.147 | 3.88 |
| Netherlands | 0.163 | 5.30 | 0.75 | 0.158 | 4.96 | 0.54 | 0.181 | 4.78 |
| Belgium | 0.164 | 5.34 | 0.44 | 0.156 | 5.01 | 0.68 | 0.199 | 5.25 |
| UK | 0.237 | 7.70 | 0.93 | 0.231 | 7.25 | 0.86 | 0.289 | 7.65 |

With the exception of South Korea and China, the results of all models are close to each other for each country.

Model 3, shows a slightly higher mortality than Models 1 and 2. This effect can be expected, since Model 3 uses estimate of cases that is effectively shifted towards a lower number of cases, for countries in the rapid growth of the epidemic. The results of Model 3 are divergent both for South Korea and China, where Model 3 returns estimate of mortality that is roughly 3 times higher than Models 1 and 2.

For most examined countries, with two exceptions, the mortality estimates obtained from all models is significantly larger than mortality for Germany. The exceptions are South Korea and Austria. South Korea by extensive testing and tracking of infections has managed to contain the epidemic and was able to quench the average number of new cases from nearly 800 in the beginning of March 2020, to less than 50 in the first ten days of April. Similarly, Austria performed extensive testing and apparently, as of April 10, managed to reduce the average number of cases from 800 at the end of March 2020, to around 300 at the time of the current study.

Models 2 and 3 are in a good agreement with data for countries that are in the growth phase of the epidemic. In particular, it is remarkable that models with parameters optimised for German data fit extremely well to data collected for USA. Apparently, they adequately describe the convolution of distribution of probability from the test with rapid, nearly exponential, growth of number of new cases.

On the other hand, these models don’t correlate well with data for countries that either successfully quenched the epidemics (such as China and South Korea), or managed to at least stabilise it and somewhat decrease the number of new cases, such as Switzerland or Austria. This is expected, since these models were developed using data from the growth phase of epidemics. One should note, that in the case of low value of r-squared, the reported results from both models are obtained mostly using formula as in Model 1 (see Eq. 9), with slightly different number of cases and deaths that are taken into account due to different weighting schemes. Therefore, their results are still meaningful, although redundant to the results of Model 1.

Interestingly, in the case of Spain and Italy, there is a significant discrepancy between r-squared computed for Model 2 and Model 3. The former is in both cases above 0.7, whereas the latter is below 0.15.

By using time average of the cases over 9 days, and including the most recent cases, Model 3 is more smooth than Model 2. Apparently, it generally fits data better in the rapid growth phase (as can be seen in Table 1, but is worse in the transition from the growth phase to the stationary, or quenched phase.

The estimates reflect large differences in testing policies between countries, see Table 3. In particular, there is a large variance in testing between the European countries. Countries with similar wealth to Germany, such as France, UK, Sweden, Netherlands or Belgium perform significantly less intensive testing relative to the state of the epidemic. Germany conducted nearly 500 tests per recorded death, whereas those countries conducted 25 to 60 tests per recorded death. The is a strong negative correlation equal −0.82 between correction factor computed for the country and a logarithm of the number of tests performed per recorded death, confirms the initial assumption that inadequate testing leads to under-reporting of infections.

**Table 3:** Summary of the state of COVID-19 epidemics in selected countries in comparison with number of tests per 1 recorded death. Data sorted by the last column. The following abbreviations are used in columns heads: Est — Estimate of number of cases obtained from the models, CF — average correction factor, DpM — deaths per 1 million population, EpM — estimated cases per 1M population, TpD — number of tests per one recorded death. Number of tests based on data collected from Worldometers.info Coronavirus service on April 12 2020.

| Country | Deaths | Cases | Est | CF | DpM | EpM | TpD |
| --- | --- | --- | --- | --- | --- | --- | --- |
| South Korea | 211 | 10 480 | 10 500 | 1.00 | 4 | 204 | 2 419 |
| Germany | 2 736 | 124 908 | 124 900 | 1.00 | 33 | 1 491 | 482 |
| Austria | 337 | 13 800 | 13 800 | 1.00 | 37 | 1 532 | 418 |
| Portugal | 470 | 15 987 | 23 000 | 1.44 | 46 | 2 260 | 346 |
| Turkey | 1 101 | 52 167 | 75 400 | 1.45 | 13 | 894 | 309 |
| Switzerland | 1 036 | 25 107 | 42 200 | 1.68 | 120 | 4 873 | 183 |
| USA | 20 463 | 526 396 | 1 196 000 | 2.27 | 62 | 3 614 | 130 |
| Sweden | 887 | 10 151 | 54 000 | 5.32 | 88 | 5 346 | 62 |
| Iran | 4 357 | 70 029 | 166 100 | 2.37 | 52 | 1 977 | 58 |
| Brazil | 1 124 | 20 727 | 78 100 | 3.77 | 5 | 367 | 56 |
| Italy | 19 468 | 152 271 | 741 400 | 4.87 | 322 | 12 264 | 49 |
| Netherlands | 2 653 | 24 571 | 129 300 | 5.26 | 155 | 7 547 | 38 |
| UK | 9 892 | 79 874 | 636 200 | 7.97 | 146 | 9 372 | 34 |
| Belgium | 3 346 | 28 018 | 177 200 | 6.33 | 289 | 15 291 | 31 |
| France | 13 851 | 130 727 | 690 500 | 5.28 | 212 | 10 579 | 24 |
| Spain | 16 606 | 163 027 | 690 800 | 4.24 | 355 | 14 776 | 21 |

## 5 Web Service

The model has been implemented as a web service that is available at covid-model.net. The main window of the service displays the summary of model results for the COVID-19 epidemics in the previous day. The view can be switched between basic mode, with summarised information, or a detailed presentation of all models. For all countries for which results of the model are available, the detailed presentation of the epidemics history is available.

Basic mode of the world table and wide mode of the country data are displayed in Figure 2. As of April 11, the estimate from at least one model was obtained for 56 countries — one model for 19, two for 37 and three for 19 countries, respectively.

**Figure 2:**
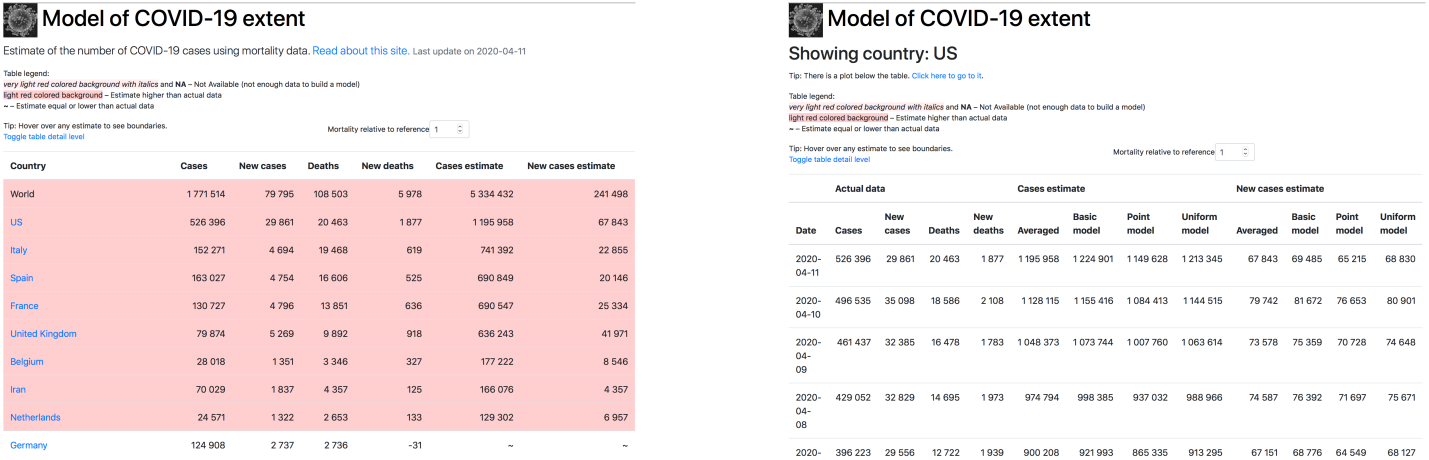
The main view of the service in the basic mode (left) and country view in wide mode (right).

The trajectory of officially recorded total cases, along with the estimate, is also available, see Figure 3. All data generated from the models is available for download from the service. The source code for the modelling is available from the authors upon request.

**Figure 3:**
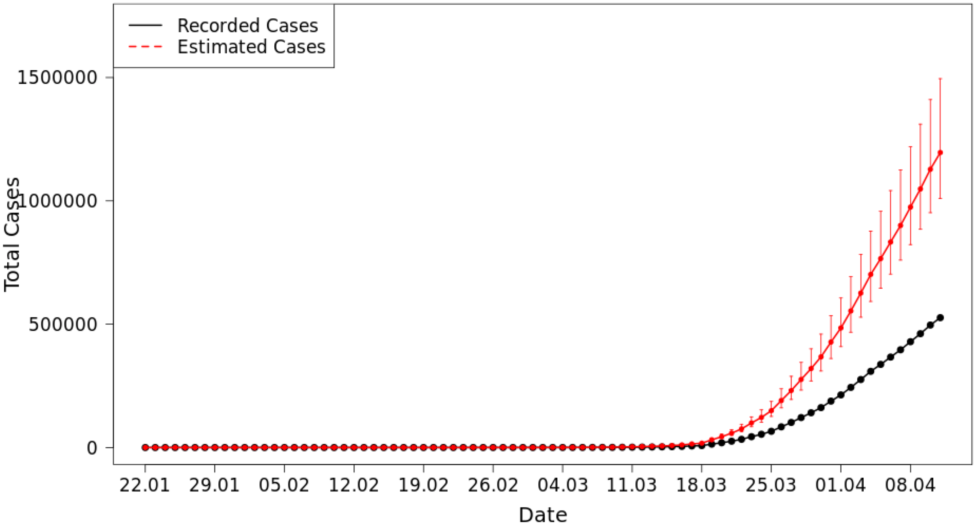
The trajectory of COVID-19 epidemics in the USA. The official data in black, the estimate in red.

## 6 Conclusions

The models proposed in the current study, while imperfect, provide a better estimate of the true extent of epidemics than the official data. The scale of discrepancy between the model and the official record is strongly correlated with the extent of testing, measured by relation of number of tests to the number of deaths attributed to COVID-19. Therefore, we suggest that the results of our model should be used as supplementary source of information about the state of epidemics for general public, researchers and public authorities.

## Data Availability

The data generated by the models is available online

https://covid-model.net

## Footnotes

1 Data read from https://www.worldometers.info/coronavirus/#countries on April 13, 2020.

2 Data read from https://www.worldometers.info/coronavirus/#countries on April 13, 2020.

